# Circulating cytokine levels and 5-year vascular recurrence after stroke: a multicenter prospective cohort study

**DOI:** 10.1101/2025.01.12.25320408

**Authors:** Lanyue Zhang, Mohamad Ali Antabi, Jana Mattar, Omar El Bounkari, Rong Fang, Karin Waegemann, Felix J. Bode, Sebastian Stösser, Peter Hermann, Thomas G. Liman, Christian H. Nolte, Benno Ikenberg, Kathleen Bernkopf, Wenzel Glanz, Daniel Janowitz, Annika Spottke, Michael Wolfgang Görtler, Silke Wunderlich, Inga Zerr, Gabor C. Petzold, Matthias Endres, Jürgen Bernhagen, Martin Dichgans, Marios K. Georgakis, DEMDAS investigators

## Abstract

**Background and Objectives:** Anti-inflammatory therapies are tested in randomized trials for secondary stroke prevention. Detecting inflammatory biomarkers that predict vascular recurrence could optimize patient selection for these trials.

**Methods:** In a multicenter prospective cohort study, we measured plasma levels of 22 inflammatory cytokines in 486 acute stroke patients (median age 68 years, 34% female, median 3 days post-stroke onset). Patients were followed for over 5 years through telephone and in-person interviews to record the occurrence of the following outcomes: (1) recurrent stroke or transient ischemic attack (TIA; primary outcome); (2) a composite of recurrent vascular events (stroke, TIA, acute coronary syndrome, hospital admission due to heart failure, and death; secondary outcome). Associations between cytokine levels and these outcomes were analyzed using Cox proportional hazards models adjusted for demographic and vascular risk factors.

**Results:** During the 5-year follow-up period, 59 patients (12.1%) experienced recurrent stroke or TIA, and 118 (24.3%) experienced recurrent vascular events. After adjustments for demographic and vascular risk factors, and correction for multiple comparisons, higher plasma levels of CD62E (adjusted Hazard Ratio [aHR] per SD increment: 1.686, 95%CI, 1.241–2.290) and MIF (aHR: 1. 627, 95%CI, 1.226–2.160) in the acute phase after stroke were statistically significantly associated with increased risk of recurrent stroke or TIA. The associations followed a dose-response pattern across quartiles of CD62E and MIF levels. Adding baseline CD62E and MIF levels to models including age, sex, vascular risk factors, and baseline C-reactive protein (CRP) levels led to significant improvements in the prediction of 5-year risk of recurrent stroke or TIA (ΔC-index 0.028 to 0.050).

**Conclusion:** Among stroke patients, higher baseline levels of CD62E and MIF improved prediction of 5-year risk of recurrent stroke or TIA on top of vascular risk factors and CRP levels. Whether assessment of these cytokines could improve patient selection for secondary prevention trials of anti-inflammatory treatments, should be explored in future studies.

## Introduction

Stroke is one of the leading causes of mortality and adult disability worldwide. In 2021, there were 93.8 million recorded prevalent strokes, accounting for 7.3 million deaths and 160.5 million disability-adjusted life-years globally.^1^ Stroke prevalence is projected to increase from 3.9% to 6.4% among the US adult population, posing a major public health burden.^2^ Despite advances in secondary prevention, stroke survivors face an alarmingly high risk of recurrent vascular events, with approximately 40% encountering a new stroke, acute coronary events, or cardiovascular death within 5 years.^3,4^ These rates point to a high residual risk that current secondary prevention strategies fail to address.^5^

A growing body of evidence supports targeting inflammation as a potential strategy for secondary stroke prevention.^6–8^ The central role of inflammation in the progression of atherosclerotic cardiovascular diseases^9^ and stroke has been well documented through experimental research^10^, genetic epidemiology^11,12^, and vascular imaging studies^13,14^. Moreover, three phase 3 trials (CANTOS^15^, LoDoCo2^16^, COLCOT^17^) provided evidence that anti-inflammatory treatment with canakinumab or colchicine reduces vascular events in patients with coronary artery disease. Aiming to translate this concept to secondary stroke prevention, the recently completed CONVINCE study which testing the non-specific anti-inflammatory drug colchicine revealed no significant reduction in recurrent vascular events among patients with a recent non-disabling non-cardioembolic stroke.^8,18^ However, notable risk reductions were observed in subgroups with atherosclerotic carotid stenosis or a history of coronary artery disease, underscoring the necessity for a more precise selection of patients who could benefit from anti-inflammatory therapies.

High-sensitivity C-reactive protein (CRP) and interleukin-6 (IL-6) are established inflammatory markers strongly associated with cardiovascular disease risk.^19,20^ Both biomarkers have been previously associated with the risk of incident^21^ and recurrent^22^ stroke. CRP has also been used to select patients for inclusion in trials testing anti-inflammatory agents for cardiovascular disease.^15,23^ However, their utility for secondary stroke prevention is debated due to high intra-patient variability, especially in the acute phase of stroke, and their lack of specificity for vascular inflammation.^24^ Moreover, both proteins are acute-phase reactants, and their levels might increase as a result of infections or systemic inflammatory conditions.^25^

In order to improve risk stratification and optimize patient selection for anti-inflammatory secondary preventive strategies, it is essential to identify and validate new biomarkers.^26^ Here, we investigated a comprehensive panel of cytokines, chemokines, and inflammatory adhesion proteins as potential markers for risk of recurrent stroke and other vascular events following an incident stroke. Specifically, we measured the baseline circulating levels of these 22 mediators shortly after stroke in a multicenter prospective cohort study of 486 patients and explored associations with risk of recurrent stroke and recurrent vascular events over a follow-up period of 5 years.

## Methods

### Study Design and Baseline Assessments

Participants were drawn from the DEMDAS study (DZNE [German Center for Neurodegenerative Diseases] - Mechanisms of Dementia After Stroke), a multicenter, prospective cohort study conducted at six tertiary stroke centers in Germany. Details on the study’s design, rationale, and baseline characteristics have been detailed elsewhere.^27,28^

In brief, DEMDAS recruited 600 patients aged 18 years or older who had experienced an acute stroke within the past five days and had no prior diagnosis of dementia. Exclusion criteria included severe comorbidities such as malignancies or end-stage renal disease, as well as the inability to provide consent. The recruitment period ran from May 2011 to January 2019. Participants underwent in-person follow-up assessments at 6-, 12-, 36-, and 60-months post-enrollment. This study adheres to the Strengthening the Reporting of Observational Studies in Epidemiology (STROBE) guidelines.^29^

Participants underwent detailed interviews using standardized questionnaires and received clinical, cognitive, and laboratory evaluations. Baseline data collection included sociodemographic details, medical and family history, and medication use. Evaluations comprised physiological and anthropometric measurements including blood pressure and body mass index (BMI), and clinical assessments using tools like the National Institutes of Health Stroke Scale (NIHSS)^30^, and stroke subtypes were classified according to the Trial of ORG 10172 in Acute Stroke Treatment (TOAST) criteria^31^.

### Standard Protocol Approvals and Patient Consent

The study was conducted in accordance with the Declaration of Helsinki and received ethical approval from institutional review boards of the participating sites. Primary ethics approval for the multicenter study was granted by the Ethics Committee of the Medical Faculty at LMU Munich (201-13) and additional approvals were obtained from the Ethics Committee of the Medical Faculty at the University of Bonn (116/13), the Ethics Committee of the University Medical Center Göttingen (21/1/12), the Ethics Committee of the Technical University of Munich (93/14 S), and the Ethics Committee of Otto von Guericke University at the Medical Faculty and University Hospital Magdeburg (66/13). The Berlin study site (Charité – Universitätsmedizin Berlin) was covered by the ethics approval of the Medical Faculty of LMU Munich, according to Section 15 of the Berlin Medical Association Professional Code of Conduct (September 2009). Written informed consent was obtained from all participants or their legal representatives.

### Blood Sample Collection and Processing

Blood samples for the current analysis were collected at baseline (within 5 days of stroke onset). The collection included one 9 mL Serum-Monovette for serum, three 9 mL EDTA-Monovettes (one specifically designated for plasma and two for DNA isolation), and one 2.5 mL Qiagen PAXGene tube for RNA-stabilized whole blood. Samples were processed immediately at each site and subsequently stored in a centralized biobank at the Institute for Stroke and Dementia Research (ISD) in Munich. Samples were centrifuged at 2,000 x g for 10 minutes at 15°C, aliquoted into 300 µL aliquots, and stored at -80°C. To minimize pre-analytical variability, rigorous quality control measures were implemented, including adherence to standard operating procedures and regular audits that have been previously described.^27^ The specimens were double-pseudonymized and recorded using a protected data integration system to maintain confidentiality and traceability.

### Cytokine Quantification and Quality Control

The Luminex® Procartaplex Multiplex Assay was used to quantify the levels of 22 cytokines in plasma baseline samples. The quantified cytokines included IP-10/CXCL10, MCP-1/CCL2, MIF, MIP-1α/CCL3, MIP-1β/CCL4, TNF-α, CD40L, CD62E/E-selectin, CD62P/P-selectin, CCL11, CSF-2, IFN-α, IFN-γ, IL-1α, IL-1β, IL-6, IL-4, IL-8/CXCL8, IL-10, IL-12p70, IL-13,and IL-17A. This panel was selected on the basis of previous evidence linking these cytokines to atherosclerosis and arterial inflammation.^32,33^ Plasma samples were thawed, mixed thoroughly, and prepared for analysis according to the manufacturer’s protocol. The Capture Bead Mix was vortexed for 30 seconds before adding 50 µL to each well of a 96-well plate. A Hand-Held Magnetic Plate Washer ensured complete beads washing. Standards were prepared using a 4-fold serial dilution to generate a standard curve for accurate quantification. Initial tests determined that the optimal dilution factor for the plasma samples was 1:4, balancing sensitivity with the assay’s dynamic range. Plasma samples (25 µL) were added to the wells containing the Capture Bead Mix and incubated on a shaker at 600 rpm for 2 hours at room temperature. The plate was analyzed using the Luminex 200 machine, and the results were processed using xPONENT v. 3.1 software (Luminex Corporation). The doublet discrimination (DD) gate was set at 7,500-25,000, with a sample volume of 50 µL, and at least 50 events per bead were recorded. A final reading volume of 120 µL per well was achieved by calibrating the device to detect at least 50 beads per analyte per sample. The concentration of the analytes was calculated using ProcartaPlex Analysis App software (Invitrogen) with a five-parameter logistic (5PL) fitting model. Data were log-transformed and standardized by batch to minimize between-batch variability, achieved by subtracting batch-specific means and dividing by batch-specific SD of ln-transformed cytokine values. Intra-assay and inter-assay coefficients of variation were monitored to ensure data reliability. Out-of-range (OOR) measurements below the detection limit, accounting for 3.9% of the data, were assigned half the lowest detectable concentration for each cytokine. Measurements exceeding 5 SD above or below the mean were excluded as potential technical errors.

### Outcome Assessment and Follow-Up

Follow-up assessments were conducted at 6, 12, 36, and up to 60 months and included clinical evaluations, imaging, and medical record reviews by study physicians. The primary outcome of the study was recurrent stroke or transient ischemic attack (TIA). Recurrent stroke was defined as the acute onset of new neurological symptoms consistent with stroke and confirmed by imaging (MRI or CT) as a new infarct or hemorrhage distinct from the initial event. TIA was defined as the acute onset of neurological symptoms that resolved within 24 hours without imaging evidence of a new infarction. The secondary composite outcome included recurrent vascular events, defined as the occurrence of any of the following after the initial stroke: myocardial infarction, angina pectoris, hospitalization due to acute or decompensated heart failure, TIA, recurrent stroke, or all-cause mortality. All outcomes were confirmed by a thorough inspection of the participants’ medical records, review of imaging studies, and clinical assessments during the in-person follow-up interviews by the study physicians. Deaths from any cause were recorded throughout the follow-up period, with the cause determined through medical record reviews and, when necessary, by contacting family members or caregivers, or accessing records from local registration offices.

### Statistical Analyses

All statistical analyses were performed using R (version 4.3.3). The following R packages were utilized: *survival* package (version 3.7-0) for survival analysis, *survminer* package (version 0.4.9) for creating Kaplan-Meier curves, *survcomp* package (version 1.52.0) for C-index calculation, and *ggplot2* (version 3.5.1) for data visualization. Baseline characteristics for continuous variables were expressed as medians with interquartile ranges (IQR) and for categorical variables as counts and percentages. For group comparisons, the Mann–Whitney U or the Kruskal-Wallis tests were used for continuous variables, and the χ² or Fisher’s exact tests were used for categorical variables.

The effects of potential determinants of cytokine levels were examined using univariable linear regression models for ln-transformed values of each cytokine. Each cytokine was modeled individually against each predictor variable to evaluate independent effects. Predictors included age, sex, BMI, low-density lipoprotein (LDL) cholesterol levels, hypertension, diabetes, history of atrial fibrillation, history of stroke, current smoking, current drinking, infarct volume, year of patient recruitment, NIHSS scores, time from symptom onset to blood collection, and CRP levels.

Cox proportional hazards regression models were employed to evaluate associations between cytokine levels and time-to-event outcomes, including primary outcomes of recurrent stroke/TIA and recurrent vascular events over a 5-year follow-up period. Ln-transformed cytokine levels were analyzed as both continuous (per SD increment) and categorical variables (split in quartiles). The associations between cytokine levels with the study outcomes were analyzed in two multivariable models adjusting for different sets of risk factors: (1) demographic factors (age, sex) and (2) demographic factors and baseline vascular factors (hypertension, diabetes, current smoking, history of stroke, history of atrial fibrillation, stroke subtypes, anticoagulants, antihypertensive medications, antiplatelet agents, statins, and LDL cholesterol).

To address the missing values in LDL cholesterol under the assumption of missing at random, we employed multiple imputation by chained equations (MICE) with predictive mean matching (PMM; five imputations, ten iterations) using the *mice* package (version 3.16.0) in R.^34^ The imputation model incorporated age, sex, hypertension, diabetes, current smoking, history of stroke, statin medication use, and BMI to predict LDL cholesterol values. All Cox regression models were executed separately within each imputed dataset. Subsequently, regression coefficients and standard errors were pooled according to Rubin’s Rules^35^ to derive combined adjusted hazard ratios (aHR), 95% confidence intervals (CI), and corresponding p-values for each adjustment model. False discovery rate (FDR) correction was applied to control for multiple comparisons in these analyses.

Sensitivity analyses were conducted to examine the robustness of the findings. Specifically, analyses were repeated after excluding patients with hemorrhagic stroke (n=12) from the initial study cohort, as well as after excluding TIA cases from the main outcomes. Kaplan-Meier curves were generated to visualize time-to-events across different cytokine quartiles, and differences between groups were compared using the log-rank test.

Model discrimination was assessed by calculating Harrell’s C-index across all imputed datasets. For each outcome, C-indices were pooled according to Rubin’s rules^35^ to account for both within-imputation and between-imputation variance. Incremental discriminative value of inflammatory biomarkers was evaluated by calculating ΔC-index between nested models, with statistical significance determined using the *cindex.comp* function. Models were constructed hierarchically, beginning with basic clinical variables (age and sex), followed by addition of either CD62E, MIF, or both biomarkers simultaneously, with further adjustment for established vascular risk factors and C-reactive protein.

## Results

### Study flowchart and baseline characteristics

DEMDAS recruited 600 participants. Due to limited reagent availability, plasma levels of 22 cytokines could only be quantified in a subset of the total study cohort. To ensure sufficient longitudinal data for robust analysis, we prioritized participants who completed at least three assessments during the 5-year follow-up (n=520). From this group, we randomly selected 491 participants for cytokine measurements, minimizing potential selection bias. Five participants were excluded because more than 80% of their cytokine measurements were classified as outliers, defined as values lower than 5 SD below the mean. As a result, 486 participants were included in the final analysis (**Figure 1**). Their baseline characteristics are summarized in **Table 1**. The baseline characteristics of included participants did not differ significantly from those of participants who were not included in the analysis **(Supplementary Table S1)**. The included participants had a median age of 68 years (IQR, 60-76 years) and 33.7% were female. **Supplementary Table S2** provides an overview of the 22 cytokines measured in this study, including their full names, abbreviations, and biological functions.

**Figure 1.**
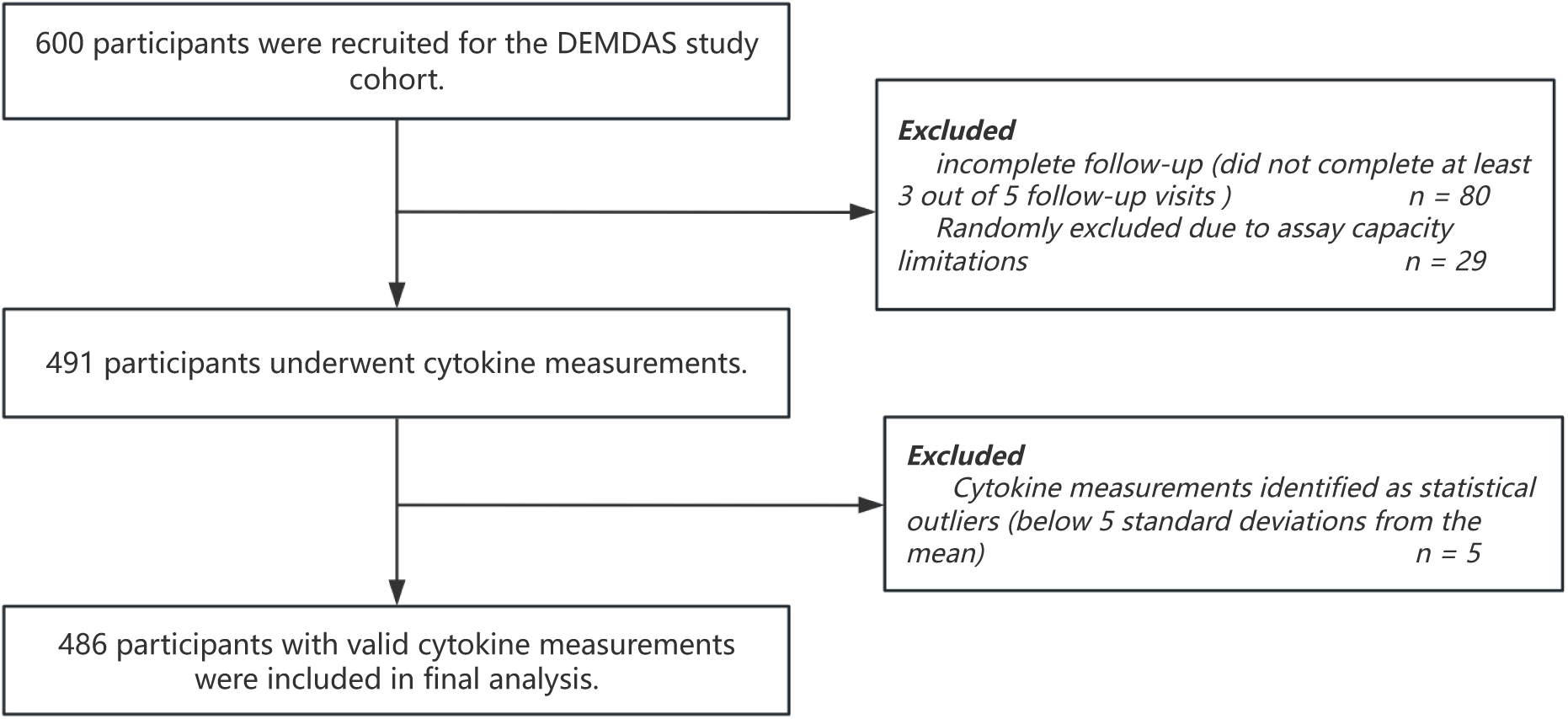
Flowchart of the participants inclusion and exclusion criteria.

**Table 1.** Baseline characteristics of participants in the DEMDAS study included in the current analysis.

| Baseline Characteristic | Overall<br>N = 486 |
| --- | --- |
| Age at recruitment (years), Median (Q1, Q3) | 68.0 (60.0, 76.0) |
| Female sex, n (%) | 164 (33.7%) |
| BMI (kg/m<sup>2</sup>), Median (Q1, Q3) | 26.5(24.3, 29.1) |
| LDL Cholesterol (mg/dL), Median (Q1, Q3) | 124.0 (101.0, 151.6) |
| C-Reactive Protein (mg/dL), Median (Q1, Q3) | 0.3 (0.1, 0.6) |
| (Missing) | 135 |
| NIHSS score at baseline, Median (Q1, Q3) | 2 (1, 5) |
| Diabetes, n (%) | 91 (19%) |
| Hypertension, n (%) | 370 (76%) |
| Atrial fibrillation, n (%) | 94 (19%) |
| Previous stroke, n (%) | 49 (10%) |
| Current smoking, n (%) | 123 (25%) |
| Current drinking, n (%) | 363 (75%) |
| Stroke classification |  |
| Ischemic stroke (TOAST subtypes), n (%) |  |
| *Large artery atherosclerosis* | 133 (27%) |
| *Cardioembolism* | 105 (22%) |
| *Small artery occlusion* | 57 (12%) |
| *Other etiology* | 25 (5.1%) |
| *Undefined etiology* | 154 (32%) |
| Hemorrhagic stroke | 12 (2.5%) |
| Stroke lesion volume (mm<sup>3</sup>), Median (Q1, Q3) | 2,344 (512, 13,048) |
| (Missing) | 18 |
| Normalized stroke lesion volume %, Median (Q1, Q3) | 0.2 (0.0, 0.8) |
| (Missing) | 22 |

Abbreviations: BMI, body mass index; LDL, low-density lipoprotein; NIHSS, National Institutes of Health Stroke Scale; TOAST, Trial of Org 10172 in Acute Stroke Treatment; IQR, interquartile range.

### Determinants of baseline cytokine levels

The distributions of ln-transformed, standardized values for the 22 quantified cytokines are illustrated in **Supplementary Figure S1**. Most cytokines showed similar patterns of variation, except for CD62E and MIF, which did not closely align with the patterns of other cytokines (**Figure 2A**, **Supplementary Table S3**). The median time between stroke onset and blood draw was 3.03 days (IQR 1.98–4.41 days) and more than 85% of the samples were fasting samples collected in the morning. Univariable linear regression analyses showed that most demographic, clinical, and technical factors had no significant associations with cytokine levels (**Figure 2B**). Most importantly, technical factors, including time from symptom onset to blood draw, as well as the year of patient recruitment (a proxy of the time that the sample remained frozen before cytokine quantification) were not associated with plasma levels of any of the cytokines (**Figure 2B**). Among vascular risk factors, we found a positive correlation between CD62E levels and BMI (Spearman’s ρ = 0.25, p-value < 0.001), as well as higher CD62E levels in patients with diabetes (**Figures 2B-D**). There were no significant differences in the levels of any of the 22 cytokines between male and female study participants (**Figure 2E, Supplementary Table S4**).

**Figure 2.**
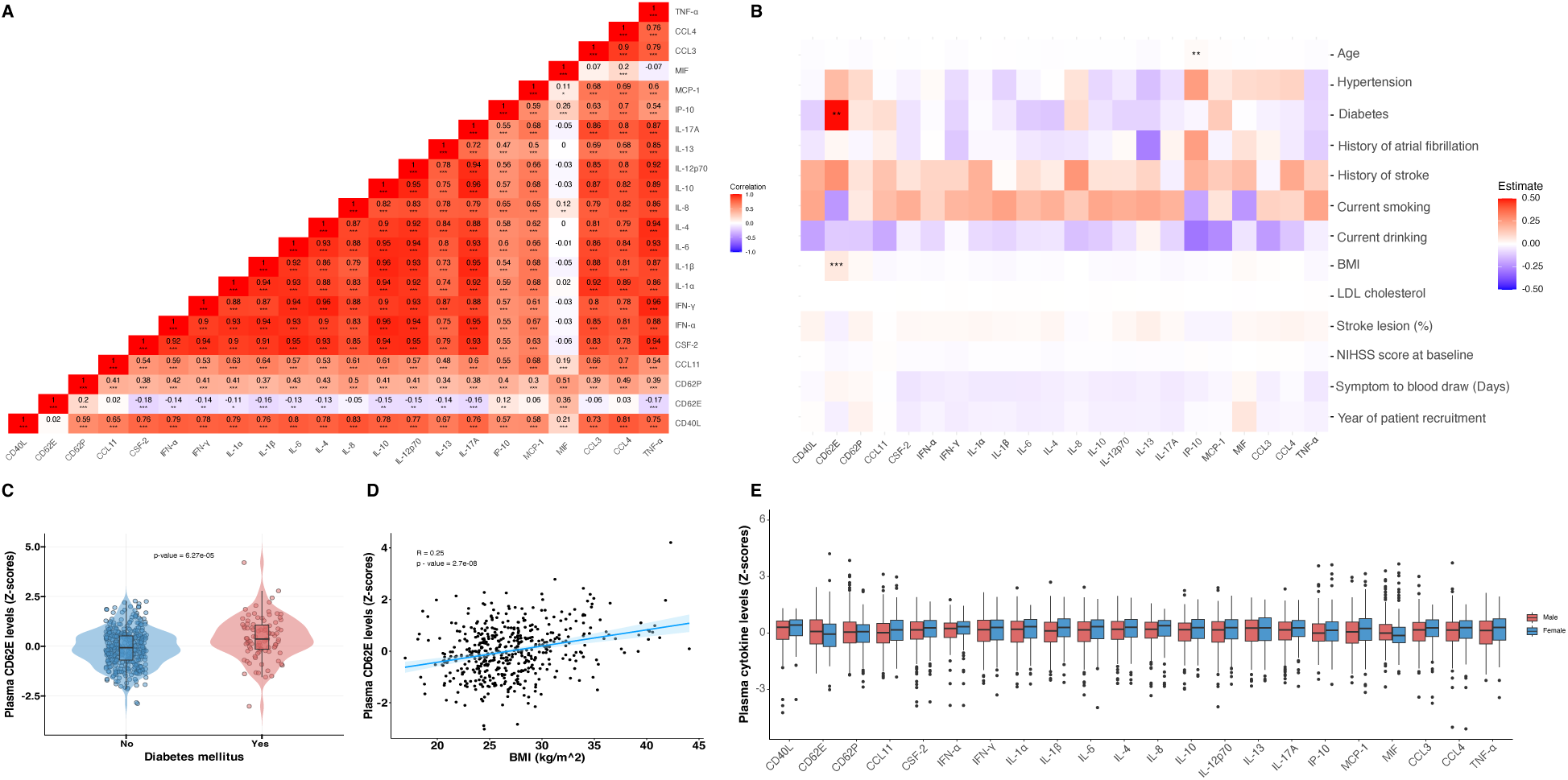
Correlations across cytokine levels and the effects of demographic, clinical, and technical factors on cytokine levels. **(A)** Heatmap illustrating the Spearman correlations across the plasma levels of 22 cytokines in the DEMDAS cohort (N = 486). The color scale ranges from blue (negative correlation) to red (positive correlation). Significance is indicated by *p-value < .05, ** p-value < .01, and *** p-value < .001, representing false discovery rate (FDR) – corrected p-values for multiple comparisons. **(B)** Heatmap illustrating the effect estimates of demographic, clinical, and technical factors on ln-transformed standardized cytokine levels, as derived from univariable linear regression analyses. Red and blue colors represent positive and negative estimates, respectively, with statistical significance marked as in (A). **(C)** Violin plot illustrating the difference in plasma CD62E levels between patients with and without diabetes mellitus. The box represents the interquartile range (IQR; 25th to 75th percentiles), with the line inside the box representing the median. **(D)** Scatter plot illustrating the positive correlation between plasma CD62E levels and BMI. **(E)** Box plot illustrating comparisons in the sex-specific distribution of cytokine levels across the DEMDAS cohort. Each box represents IQR, covering the 25th to 75th percentiles of the data, with the line inside the box indicating the median value. The whiskers extend to the most extreme data points within 1.5 times the IQR. Abbreviations: BMI, body mass index, LDL, low density lipoprotein; NIHSS, National Institutes of Health Stroke Scale; IQR, interquartile range; FDR, false discovery rate.

### Associations between cytokine levels and recurrent events

During the 5-year follow-up period, 59 (12.1%) participants experienced a recurrent stroke or TIA, and 118 (24.3%) participants experienced a composite of recurrent vascular events, including stroke, TIA, acute coronary syndrome, acute or decompensated heart failure, or death. **Figure 3** illustrates the aHR for the associations between the levels of the 22 cytokines (per SD increment in ln-transformed values) and recurrent stroke or TIA (**Figure 3A, Supplementary Table S5**) as well as recurrent vascular events (**Figure 3B, Supplementary Table S6**), derived from Cox regression analyses adjusted for age, sex, and vascular risk factors. Plasma levels of CD62E and MIF were associated with a higher risk of recurrent stroke or TIA over the 5-year follow-up period (FDR-adjusted p-value = 0.009) (**Figure 3A, Supplementary Table S5**). These associations remained significant when adjusting for age and sex (aHR for CD62E: 1.642 [95% CI, 1.244–2.168]; aHR for MIF: 1.586 [95% CI, 1.203–2.091]) and also when additionally adjusting for vascular risk factors (hypertension, diabetes, current smoking, history of stroke, history of atrial fibrillation, stroke subtypes, anticoagulants, antihypertensive medications, antiplatelet agents, statins, and low-density lipoprotein levels; aHR for CD62E: 1.686 [95% CI, 1.241–2.290]; aHR for MIF: 1.627 [95% CI, 1.226–2.160]). Although the associations between CD62E and MIF and recurrent vascular events also met the nominal threshold of p-value < 0.05, they were not significant after FDR correction (**Figure 3B**, **Supplementary Table S6**). Sensitivity analyses, excluding patients with hemorrhagic stroke (n = 12) at baseline (**Supplementary Figure S2, Supplementary Tables S7-S8**) and excluding TIA from the outcomes of interest (**Supplementary Figure S3, Supplementary Tables S9-S10**) yielded similar with consistent effects for both CD62E and MIF.

**Figure 3.**
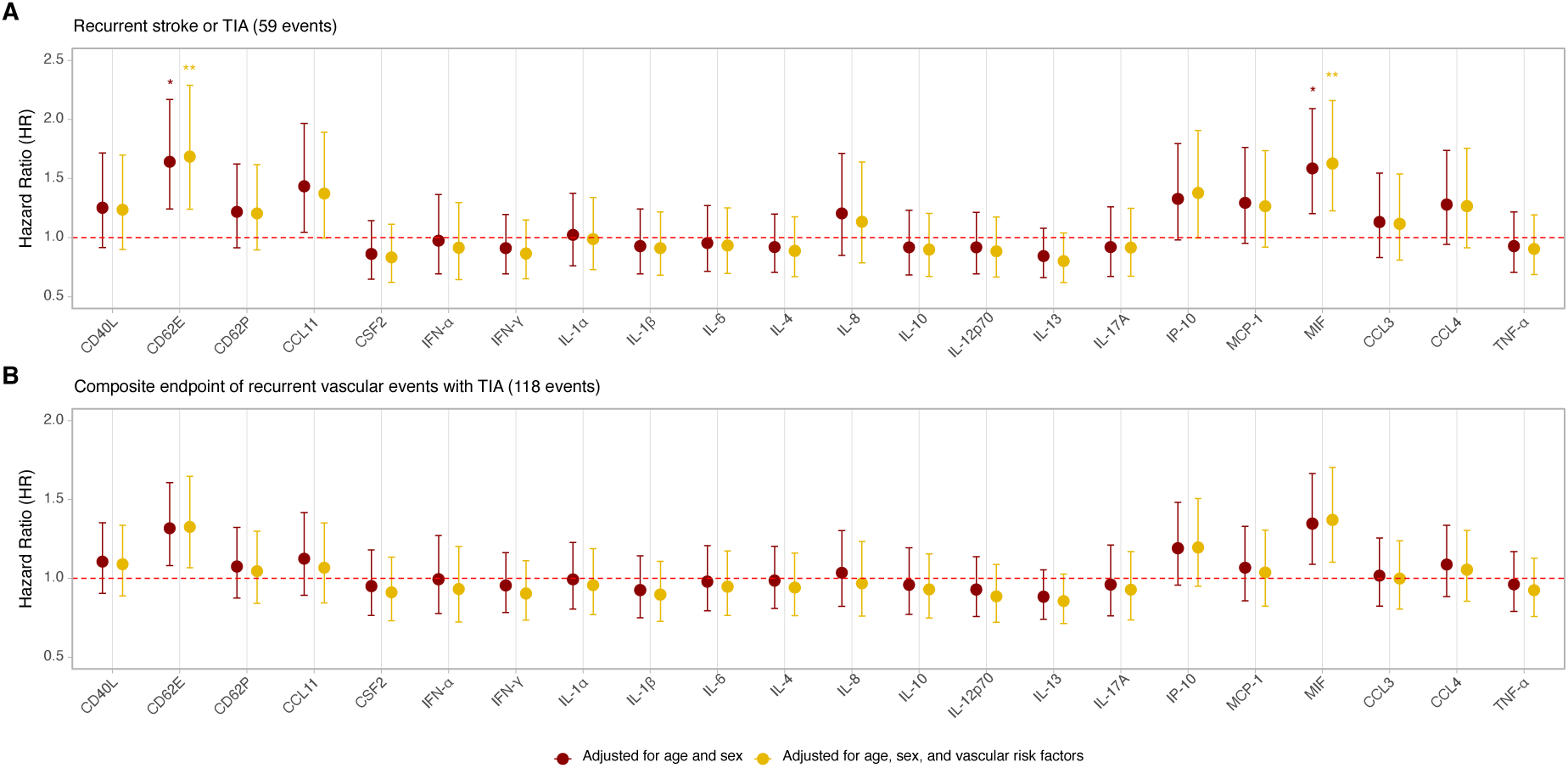
Associations of cytokine levels with recurrent stroke or transient ischemic attack (TIA) and recurrent vascular events (stroke, TIA, acute coronary syndrome, new onset of heart failure, or death) over the 5-year follow-up period. Forest plots of hazard ratios (HR) for the associations of circulating baseline cytokine levels (per SD increment in ln-transformed values) with **(A)** recurrent stroke or TIA (n = 59) and **(B)** the composite endpoint of recurrent vascular events (n = 118) over the 5-year follow-up period. The HR in the first model (red) are adjusted for age and sex, while those in the second model (yellow) are further adjusted for vascular risk factors, including hypertension, diabetes, current smoking, history of stroke, history of atrial fibrillation, stroke subtypes, anticoagulants, antihypertensive medications, antiplatelet agents, statins, and low-density lipoprotein levels. Vertical error bars indicate the 95% CI for each HR. Significance is indicated by * p-value < .05, representing FDR-corrected p-values for multiple comparisons. Abbreviations: TIA, transient ischemic attack; HR, hazard ratio; CI, confidence interval; FDR, false discovery rate.

When analyzing by quartiles of the baseline CD62E and MIF distributions, dose dependent increases in the risk of recurrent stroke or TIA were observed for both proteins (**Figure 4**). Compared with the first quartile (Q1), patients in the highest quartiles of CD62E and MIF distributions (Q4) were at statistically significantly higher risk of recurrent stroke or TIA over the 5-year follow-up period (aHR for CD62E Q4 vs. Q1: 3.070 [95% CI, 1.384–6.812]; aHR for MIF Q4 vs. Q1: 2.636 [95% CI, 1.190–5.837]). These effects remained robust after adjusting for age, sex, and vascular risk factors, as well as when excluding TIA from the definition of the outcome (**Supplementary Figure S4**). For CD62E, similar albeit weaker associations were noted for the composite endpoint of recurrent vascular events (**Supplementary Figure S5**).

**Figure 4.**
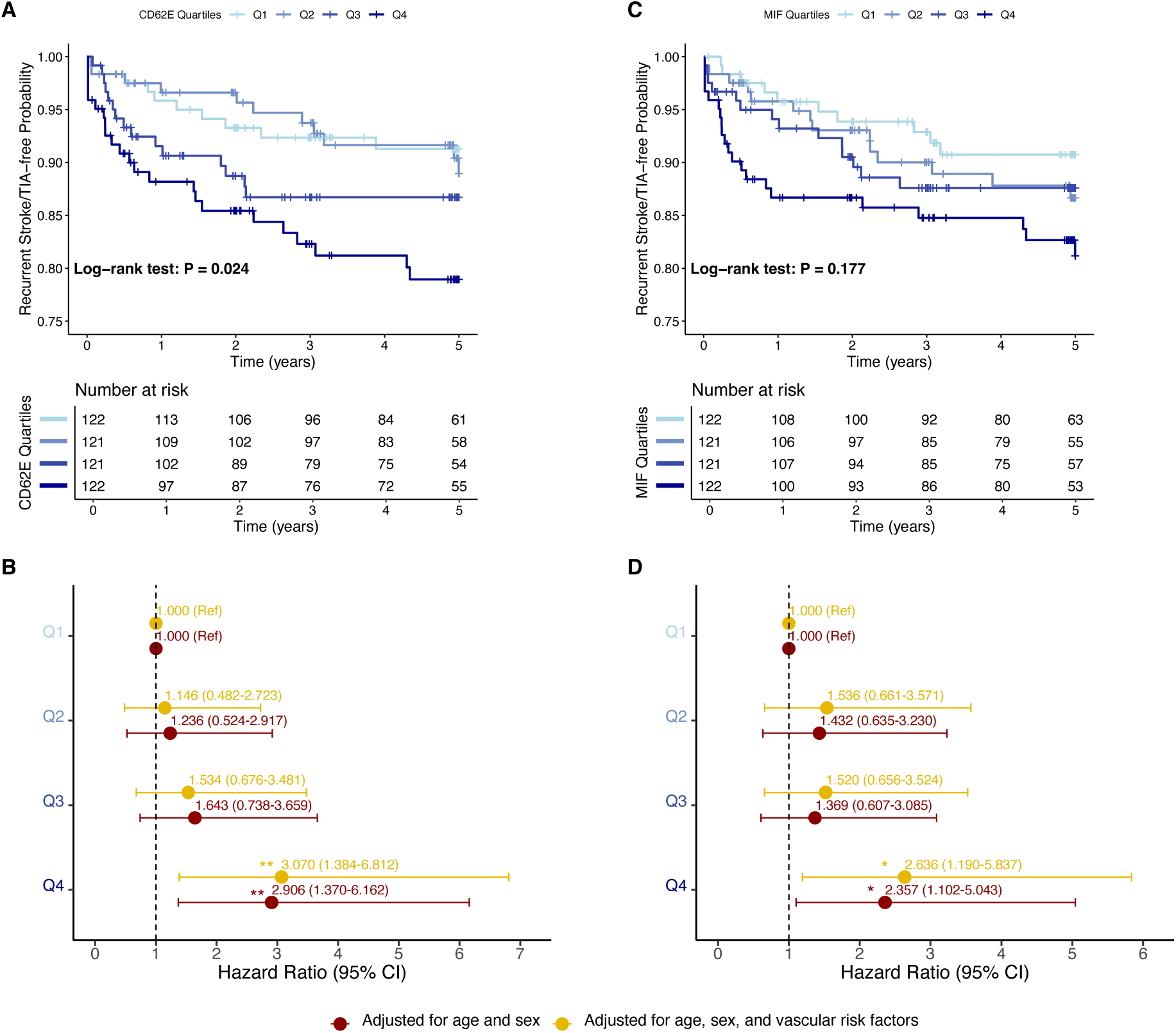
Dose-response associations of baseline CD62E and MIF levels with 5-year risk of recurrent stroke or transient ischemic attack (TIA). **(A)** Kaplan-Meier 5-year recurrent stroke- or TIA-free survival curves for DEMDAS participants across quartiles (Q1 to Q4) of baseline circulating CD62E levels. **(B)** Forest plot of adjusted HR for recurrent stroke or TIA across quartiles of baseline circulating CD62E levels. **(C)** Kaplan-Meier 5-year recurrent stroke- or TIA-free survival curves for DEMDAS participants across quartiles (Q1 to Q4) of baseline circulating MIF levels. **(D)** Forest plot of adjusted HR for recurrent stroke or TIA across quartiles of baseline circulating MIF levels. In panels (B) and (D), the HR in the first model (red) are adjusted for age and sex, while the HR in the second model (yellow) are further adjusted for vascular risk factors, including hypertension, diabetes, current smoking, history of stroke, history of atrial fibrillation, stroke subtypes, anticoagulants, antihypertensive medications, antiplatelet agents, statins, and low-density lipoprotein levels. Horizontal lines represent the 95% CI for each HR. Statistical significance is indicated by *p-value < .05, **p-value < .01. Abbreviations: TIA, transient ischemic attack; HR, hazard ratio; CI, confidence interval.

### Predictive performance of CD62E and MIF for recurrent events

As a final step, we assessed whether adding baseline CD62E and MIF levels to models including baseline demographic and vascular risk factors, as well as CRP levels, could improve the prediction of recurrent stroke or TIA (**Table 2**). The addition of baseline CD62E and MIF levels improved the C-index of Cox regression models for 5-year risk of recurrent stroke or TIA on top of age and sex (ΔC-index = 0.050, p-value = 0.046), age, sex, and vascular risk factors (ΔC-index = 0.033, p-value = 0.026), as well as age, sex, vascular risk factors, and baseline CRP levels (ΔC-index = 0.028, p-value = 0.032). Similar improvements were observed when analyzing 5-year risk of recurrent stroke, excluding TIA cases (**Supplementary Table S11**).

**Table 2.**
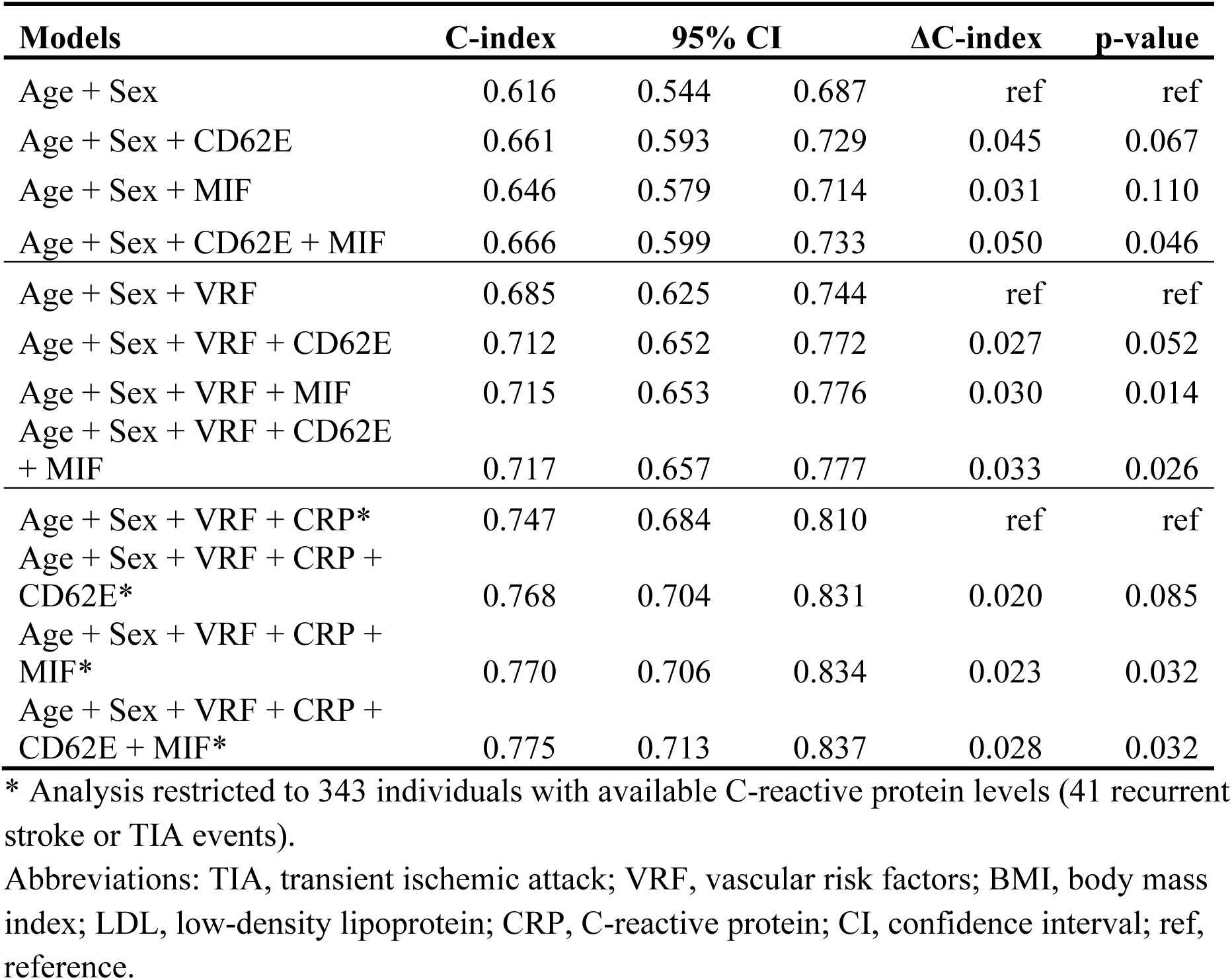
C-index comparisons of predictive models for 5-year risk of recurrent stroke or transient ischemic attack (TIA), incorporating different combinations of age, sex, vascular risk factors (VRF; hypertension, diabetes, current smoking, history of stroke, history of atrial fibrillation, stroke subtypes, anticoagulants, antihypertensive medications, antiplatelet agents, statins, and LDL cholesterol), and baseline circulating levels of C-reactive protein (CRP), CD62E, and MIF.

## Discussion

In this multicenter prospective cohort study of 486 acute stroke patients, we aimed to comprehensively evaluate the predictive value of a panel of 22 cytokines for risk of vascular recurrence. We found that higher baseline levels of CD62E and MIF were associated with an increased risk of recurrent stroke or TIA over a 5-year follow-up period. The associations were robust to adjustments for baseline demographic and vascular risk factors and followed dose-response patterns. Adding baseline CD62E and MIF levels on top of age, sex, vascular risk factors, and baseline CRP levels led to significant improvements in risk discrimination for recurrent stroke or TIA (ΔC-index: 0.028 to 0.050).

Our findings may have important implications for risk stratification in stroke patients, particularly in selecting candidates for anti-inflammatory treatments. In line with the CANTOS trial^15^, the ongoing ZEUS trial^23^, which tests an anti-IL-6 antibody in a broad population with atherosclerosis (including atherosclerotic stroke), uses a CRP threshold of >2 mg/dL to select patients. Similarly, the CASPER trial, which tests colchicine in ischemic stroke patients, restricts enrollment to those with CRP levels >2 mg/dL.^36^ While a meta-analysis supports an association between CRP levels and stroke recurrence,^22^ CRP is not specific for arterial inflammation, and transient increases in the acute phase after stroke could limit its utility for risk stratification at that critical time window.^37^ Here, we provide evidence that two other inflammatory mediators, CD62E and MIF, may offer improved prediction of recurrent stroke beyond traditional risk factors and CRP levels. Notably, the point estimates for the associations between CD62E and MIF and recurrent stroke risk were larger than previously reported for CRP (CD62E: HR for Q4 vs. Q1 3.070; MIF: HR for Q4 vs. Q1: 2.636; CRP: HR for Q4 vs. Q1 1.40 when adjusting for age and sex).^22^ Furthermore, in the largest meta-analysis to date, the association of CRP with recurrent stroke was barely significant following adjustments for vascular risk factors, whereas CD62E and MIF maintained significant associations with recurrent stroke risk. While CD62E and MIF levels are known to be elevated in certain infections, such as infective endocarditis and brucellosis,^38–40^ their associations with recurrent stroke in our study remained significant after adjusting for CRP, suggesting their predictive value may extend beyond infection-related inflammation. Whether these biomarkers could aid in selecting patients for anti-inflammatory treatments requires further investigation in external cohorts and in *post hoc* analysis of randomized trial data. It is noteworthy, however, that despite the significant association of the highest quartile (Q4) of MIF with recurrent stroke/TIA in adjusted Cox models, the Kaplan–Meier analysis did not demonstrate statistically significant differences among MIF quartiles (log-rank P = 0.177). This discrepancy could reflect limited statistical power due to the moderate sample size and relatively lower event rate, emphasizing the need for validation studies with larger cohorts to clarify the clinical utility of MIF measurement.

Both CD62E and MIF have been extensively linked to the pathophysiology of vascular disease. CD62E, an adhesion molecule also known as E-selectin, mediates leukocyte adhesion to the endothelium, a critical step in the recruitment of immune cells to the subendothelial space and the initiation of arterial inflammation.^41^ As inflammation progresses, CD62E undergoes increased shedding, making it detectable in plasma and reflecting its upregulation. This shedding process, mediated by metalloproteinases such as ADAM17, has been shown to regulate adhesion molecule function and contribute to inflammatory signaling pathways.^42–45^ Mouse studies have shown that targeting E-selectin reduces atherosclerotic lesions and improves heart function.^46,47^ In acute coronary syndrome, elevated CD62E levels reflect endothelial activation, promoting leukocyte adhesion—a hallmark of inflammation and vascular dysfunction.^47,48^ While studies directly linking CD62E to stroke recurrence are limited, it has been identified as a predictor of poor 3-month outcomes in stroke patients, including functional impairment.^49^ Experimental models further demonstrate that blocking E-selectin improves cerebral blood flow and reduces infarct size, highlighting its therapeutic potential.^50^ Additionally, MR analyses have associated genetically proxied CD62E with adverse post-stroke outcomes, reinforcing its role in cerebrovascular disease.^51^ Similarly, MIF—a pro-inflammatory cytokine and atypical chemokine^52^, drives the atherogenic recruitment of monocytes, neutrophils, T cells, and platelets to atherosclerotic plaques and has been implicated in plaque instability and rupture.^53–56^ Targeting MIF has been shown to reduce atherosclerosis in experimental mouse models.^54,57–60^ A study of 469 stroke patients with a shorter follow-up (12 months) also found significant associations between baseline MIF levels and risk of recurrent stroke.^61^ In another study of stroke survivors, elevated MIF levels in the acute phase after stroke were linked to poorer stroke outcomes.^62^

Our study has several limitations. First, cytokine levels were measured only once, shortly after stroke onset, without assessing changes over time. Serial measurements could provide insights into the temporal dynamics of inflammatory markers and their relationship with recurrence. Second, despite our efforts, the DEMDAS cohort had a relatively high attrition rate of about 15% over the 5-year follow-up period, potentially leading to the selection of less severely affected patients and an underestimation of recurrence rates. While this loss to follow-up could introduce selection bias, baseline demographic and clinical characteristics of patients included in and excluded from our analyses were similar. Third, the comprehensive follow-up protocol, which included serial imaging and neuropsychological assessments, led to the preferential recruitment of patients with milder strokes.^27,63^ Although this might limit the generalizability of our findings to a broader stroke population, these patients may represent a group that could benefit from more aggressive secondary prevention including anti-inflammatory therapies. Fourth, the relatively small number of recurrent vascular events — likely due to the mildly affected stroke population — may have reduced statistical power to detect significant associations with other promising cytokines after multiple testing corrections. Fifth, CRP levels were missing in a substantial proportion of our study population (29%), further limiting statistical power when comparing CD62E and MIF levels with CRP. Sixth, because of limited data on the exact causes of death, our definition of recurrent vascular events included all-cause mortality rather than only vascular death, deviating from the standard definition of major adverse cardiovascular events.^64^ However, since cardiovascular disease is the most common cause of death among stroke survivors^65^ and since there were only 46 deaths over the 5-year follow-up period, we do not expect this to have introduced substantial misclassification bias.

In conclusion, our study demonstrates that elevated plasma levels of CD62E and MIF shortly after acute stroke are associated with a higher 5-year risk of recurrent stroke or TIA, offering improved predictive value on top of traditional risk factors and CRP levels. However, these findings are preliminary and need to be confirmed through larger studies to establish their clinical relevance. If validated in future studies, assessing these cytokines could enhance risk stratification for stroke patients and inform patient selection for randomized trials of anti-inflammatory agents.

## Data Availability

All data produced in the present study are available upon reasonable request to the authors.

## Acknowledgements

We thank all patients for their study participation. We appreciate the support of Barbara Klapacz and Regina Altmann with data management and would like to thank Tatjana Wittenberg, Marion Sengewald, Sara Schmidt, Sandra Becker, Julia Schütte-Schmidt, Franziska Schulze, Christine Chahli, Esther D’Andrade, and Annette Eder for assistance with recruitment and data acquisition.

## References

1. Feigin VL, Abate MD, Abate YH, et al. Global, regional, and national burden of stroke and its risk factors, 1990–2021: a systematic analysis for the Global Burden of Disease Study 2021. The Lancet Neurology. 2024;23:973–1003.

2. Joynt Maddox KE, Elkind MSV, Aparicio HJ, et al. Forecasting the Burden of Cardiovascular Disease and Stroke in the United States Through 2050—Prevalence of Risk Factors and Disease: A Presidential Advisory From the American Heart Association. Circulation. 2024;150:e65–e88.

3. Flach C, Muruet W, Wolfe CDA, et al. Risk and Secondary Prevention of Stroke Recurrence. Stroke. 2020;51:2435–2444.

4. Chen Y, Wright N, Guo Y, et al. Mortality and recurrent vascular events after first incident stroke: a 9-year community-based study of 0·5 million Chinese adults. The Lancet Global Health. 2020;8:e580–e590.

5. Hankey GJ. Secondary stroke prevention. The Lancet Neurology. 2014;13:178–194.

6. Ajala ON, Everett BM. Targeting Inflammation to Reduce Residual Cardiovascular Risk. Curr Atheroscler Rep. 2020;22:66.

7. Zietz A, Gorey S, Kelly PJ, Katan M, McCabe JJ. Targeting inflammation to reduce recurrent stroke. International Journal of Stroke. 2024;19:379–387.

8. Kelly P, Lemmens R, Weimar C, et al. Long-term colchicine for the prevention of vascular recurrent events in non-cardioembolic stroke (CONVINCE): a randomised controlled trial. The Lancet. 2024;404:125–133.

9. Hansson GK, Hansson GK, Hansson GK, Hansson GK. Inflammation, Atherosclerosis, and Coronary Artery Disease. N Engl J Med. 2005;352:1685–1695.

10. Cao J, Roth S, Zhang S, et al. DNA-sensing inflammasomes cause recurrent atherosclerotic stroke. Nature. Epub 2024 Aug 7.:1–9.

11. Coveney S, Murphy S, Belton O, et al. Inflammatory cytokines, high-sensitivity C-reactive protein, and risk of one-year vascular events, death, and poor functional outcome after stroke and transient ischemic attack. International Journal of Stroke. 2022;17:163–171.

12. McPherson R, Davies RW, McPherson R, et al. Inflammation and Coronary Artery Disease: Insights From Genetic Studies. Canadian Journal of Cardiology. 2012;28:662– 666.

13. Marnane M, Merwick A, Sheehan OC, et al. Carotid plaque inflammation on ^18^F-fluorodeoxyglucose positron emission tomography predicts early stroke recurrence. Annals of Neurology. 2012;71:709–718.

14. McCabe JJ, Camps-Renom P, Giannotti N, et al. Carotid Plaque Inflammation Imaged by PET and Prediction of Recurrent Stroke at 5 Years. Neurology. 2021;97:e2282– e2291.

15. Ridker PM, Everett BM, Thuren T, et al. Antiinflammatory Therapy with Canakinumab for Atherosclerotic Disease. N Engl J Med. 2017;377:1119–1131.

16. Nidorf SM, Fiolet ATL, Mosterd A, et al. Colchicine in Patients with Chronic Coronary Disease. N Engl J Med. 2020;383:1838–1847.

17. Tardif J-C, Kouz S, Waters DD, et al. Efficacy and Safety of Low-Dose Colchicine after Myocardial Infarction. N Engl J Med. 2019;381:2497–2505.

18. Fiolet ATL, Poorthuis MHF, Opstal TSJ, et al. Colchicine for secondary prevention of ischaemic stroke and atherosclerotic events: a meta-analysis of randomised trials. eClinicalMedicine. 2024;76:102835.

19. Burger PM, Koudstaal S, Mosterd A, et al. C-Reactive Protein and Risk of Incident Heart Failure in Patients With Cardiovascular Disease. Journal of the American College of Cardiology. 2023;82:414–426.

20. Scirica BM, Morrow DA. The Verdict Is Still Out. Circulation. 2006;113:2128–2151.

21. Collaboration TERF. C-reactive protein concentration and risk of coronary heart disease, stroke, and mortality: an individual participant meta-analysis. The Lancet. 2010;375:132–140.

22. McCabe JJ, Walsh C, Gorey S, et al. C-Reactive Protein, Interleukin-6, and Vascular Recurrence After Stroke: An Individual Participant Data Meta-Analysis. Stroke. 2023;54:1289–1299.

23. Novo Nordisk A/S. ZEUS - A research study to look at how Ziltivekimab works compared to placebo in people with cardiovascular disease, chronic kidney disease and inflammation (ZEUS). ClinicalTrials.gov identifier: NCT05021835. Updated October 15, 2024. Accessed [cited 2024 Nov 12]. Available from: https://clinicaltrials.gov/study/NCT05021835

24. Ridker PM, Danielson E, Fonseca FAH, et al. Rosuvastatin to Prevent Vascular Events in Men and Women with Elevated C-Reactive Protein. N Engl J Med. 2008;359:2195– 2207.

25. O’Donnell MA. Yaron Fuchs: Exploring the mysterious mixture of life and death. J Cell Biol. 2017;216:2600–2601.

26. Lindblad M, Unbeck M, Nilsson L, Schildmeijer K, Ekstedt M. Identifying no-harm incidents in home healthcare: a cohort study using trigger tool methodology. BMC Health Serv Res. 2020;20:289.

27. Georgakis MK, Fang R, Düring M, et al. Cerebral small vessel disease burden and cognitive and functional outcomes after stroke: A multicenter prospective cohort study. Alzheimer’s & Dementia. 2023;19:1152–1163.

28. von Rennenberg R, Nolte CH, Liman TG, et al. High-Sensitivity Cardiac Troponin T and Cognitive Function Over 12 Months After Stroke—Results of the DEMDAS Study. Journal of the American Heart Association. 2024;13:e033439.

29. von Elm E, Altman DG, Egger M, Pocock SJ, Gøtzsche PC, Vandenbroucke JP. The Strengthening the Reporting of Observational Studies in Epidemiology (STROBE) statement: guidelines for reporting observational studies. The Lancet. 2007;370:1453– 1457.

30. Adams HP, Davis PH, Leira EC, et al. Baseline NIH Stroke Scale score strongly predicts outcome after stroke. Neurology. 1999;53:126–126.

31. Adams HP, Bendixen BH, Kappelle LJ, et al. Classification of subtype of acute ischemic stroke. Definitions for use in a multicenter clinical trial. TOAST. Trial of Org 10172 in Acute Stroke Treatment. Stroke. 1993;24:35–41.

32. Sterpetti AV. Inflammatory Cytokines and Atherosclerotic Plaque Progression. Therapeutic Implications. Curr Atheroscler Rep. 2020;22:75.

33. Kong P, Cui Z-Y, Huang X-F, Zhang D-D, Guo R-J, Han M. Inflammation and atherosclerosis: signaling pathways and therapeutic intervention. Signal Transduct Target Ther. 2022;7:131.

34. Buuren S van, Groothuis-Oudshoorn K. mice: Multivariate Imputation by Chained Equations in*R*. J Stat Soft. 2011;45:1–67.

35. Marshall A, Altman DG, Holder RL, Royston P. Combining estimates of interest in prognostic modelling studies after multiple imputation: current practice and guidelines. BMC Med Res Methodol. 2009;9:57.

36. Australian New Zealand Clinical Trials Registry (ANZCTR). The effect of Colchicine on Cardiovascular Outcomes in Stroke Study (The CASPER Study). ACTRN12621001408875. Updated October 20, 2021. Accessed [cited 2024 Nov 12];Available from: https://www.anzctr.org.au/Trial/Registration/TrialReview.aspx?id=382337&showOriginal=true&isReview=true.

37. Winbeck K, Poppert H, Etgen T, Conrad B, Sander D. Prognostic Relevance of Early Serial C-Reactive Protein Measurements After First Ischemic Stroke. Stroke. 2002;33:2459–2464.

38. Sumaiya K, Langford D, Natarajaseenivasan K, et al. Macrophage migration inhibitory factor (MIF): A multifaceted cytokine regulated by genetic and physiological strategies. Pharmacology & Therapeutics. 2022;233:108024.

39. Ferrero MC, Bregante J, Delpino MV, et al. Proinflammatory response of human endothelial cells to Brucella infection. Microbes and Infection. 2011;13:852–861.

40. Snipsøyr MG, Ludvigsen M, Petersen E, et al. A systematic review of biomarkers in the diagnosis of infective endocarditis. International Journal of Cardiology. 2016;202:564– 570.

41. Munro JM. Endothelial-leukocyte adhesive interactions in inflammatory diseases. Eur Heart J. 1993;14 Suppl K:72–77.

42. Reiss K, Ludwig A, Saftig P. Breaking up the tie: Disintegrin-like metalloproteinases as regulators of cell migration in inflammation and invasion. Pharmacology & Therapeutics. 2006;111:985–1006.

43. Garton KJ, Gough PJ, Philalay J, et al. Stimulated Shedding of Vascular Cell Adhesion Molecule 1 (VCAM-1) Is Mediated by Tumor Necrosis Factor-α-converting Enzyme (ADAM 17). Journal of Biological Chemistry. 2003;278:37459–37464.

44. Thomson AW, Satoh S, Nüssler AK, et al. Circulating intercellular adhesion molecule-1 (ICAM-1) in autoimmune liver disease and evidence for the production of ICAM-1 by cytokine-stimulated human hepatocytes. Clinical and Experimental Immunology. 2008;95:83–90.

45. Huber AR, Kunkel SL, Todd RF, et al. Regulation of Rransendothelial Neutrophil Migration by Endogenous Interleukin-8. Science. 1991;254:99–102.

46. Tsoref O, Tyomkin D, Amit U, et al. E-selectin-targeted copolymer reduces atherosclerotic lesions, adverse cardiac remodeling, and dysfunction. Journal of Controlled Release. 2018;288:136–147.

47. Serebruany VL, Glassman AH, Malinin AI, et al. Enhanced platelet/endothelial activation in depressed patients with acute coronary syndromes. Blood Coagulation & Fibrinolysis. 2003;14:563–567.

48. Sarrafzadegan N, Sadeghi M, Ghaffarpasand F, et al. Interleukin-6 and E-selectin in acute coronary syndromes and stable angina pectoris. Herz. 2012;37:926–930.

49. Richard S, Lagerstedt L, Burkhard PR, Debouverie M, Turck N, Sanchez J-C. E-selectin and vascular cell adhesion molecule-1 as biomarkers of 3-month outcome in cerebrovascular diseases. J Inflamm. 2015;12:61.

50. Huang J, Choudhri TF, Winfree CJ, et al. Postischemic Cerebrovascular E-Selectin Expression Mediates Tissue Injury in Murine Stroke. Stroke. 2000;31:3047–3053.

51. Wang M, Zhang Z, Liu D, et al. Soluble adhesion molecules and functional outcome after ischemic stroke: A Mendelian randomization study. Journal of Stroke and Cerebrovascular Diseases. 2023;32:107136.

52. Calandra T, Roger T, Calandra T, Roger T. Macrophage migration inhibitory factor: a regulator of innate immunity. Nat Rev Immunol. 2003;3:791–800.

53. Schober A, Bernhagen J, Thiele M, et al. Stabilization of Atherosclerotic Plaques by Blockade of Macrophage Migration Inhibitory Factor After Vascular Injury in Apolipoprotein E–Deficient Mice. Circulation. 2004;109:380–385.

54. Bernhagen J, Krohn R, Lue H, et al. MIF is a noncognate ligand of CXC chemokine receptors in inflammatory and atherogenic cell recruitment. Nat Med. 2007;13:587–596.

55. Asare Y, Schmitt M, Bernhagen J. The vascular biology of macrophage migration inhibitory factor (MIF). Thromb Haemost. 2013;109:391–398.

56. Wirtz TH, Tillmann S, Strüßmann T, et al. Platelet-derived MIF: A novel platelet chemokine with distinct recruitment properties. Atherosclerosis. 2015;239:1–10.

57. Burger-Kentischer A, Göbel H, Kleemann R, et al. Reduction of the aortic inflammatory response in spontaneous atherosclerosis by blockade of macrophage migration inhibitory factor (MIF). Atherosclerosis. 2006;184:28–38.

58. Verschuren L, Kooistra T, Bernhagen J, et al. MIF deficiency reduces chronic inflammation in white adipose tissue and impairs the development of insulin resistance, glucose intolerance, and associated atherosclerotic disease. Circulation Research. 2009;105:99–107.

59. Sinitski D, Kontos C, Krammer C, et al. Macrophage Migration Inhibitory Factor (MIF)-Based Therapeutic Concepts in Atherosclerosis and Inflammation. Thromb Haemost. 2019;119:553–566.

60. Kontos C, El Bounkari O, Krammer C, et al. Designed CXCR4 mimic acts as a soluble chemokine receptor that blocks atherogenic inflammation by agonist-specific targeting. Nat Commun. 2020;11:5981.

61. Wang G, Li C, Liu Y, et al. Macrophage Migration Inhibitory Factor Levels Correlate with Stroke Recurrence in Patients with Ischemic Stroke. Neurotox Res. 2019;36:1–11.

62. Wang C-W, Ma P-J, Wang Y-Y, et al. Serum level of macrophage migration inhibitory factor predicts severity and prognosis in patients with ischemic stroke. Cytokine. 2019;115:8–12.

63. Fang R, Duering M, Bode FJ, et al. Risk factors and clinical significance of post-stroke incident ischemic lesions. Alzheimer’s & Dementia [online serial]. n/a. Accessed at: https://alz-journals.onlinelibrary.wiley.com/doi/10.1002/alz.14274. Accessed November 12, 2024.

64. Bosco E, Hsueh L, McConeghy KW, Gravenstein S, Saade E. Major adverse cardiovascular event definitions used in observational analysis of administrative databases: a systematic review. BMC Med Res Methodol. 2021;21:241.

65. Brønnum-Hansen H, Davidsen M, Thorvaldsen P. Long-Term Survival and Causes of Death After Stroke. Stroke. American Heart Association; 2001;32:2131–2136.

