## Supplemental Figures S1-S5 for "Circulating cytokine levels and 5-year vascular recurrence after stroke: a multicenter prospective cohort study"

[#] Names and affiliations of DEMDAS investigators are listed in the supplement (**Table S12**)

**Corresponding author**

Marios K. Georgakis, MD, PhD
Institute for Stroke and Dementia Research
Ludwig-Maximilians-University Hospital
Feodor-Lynen-Str. 17, 81377 Munich, Germany


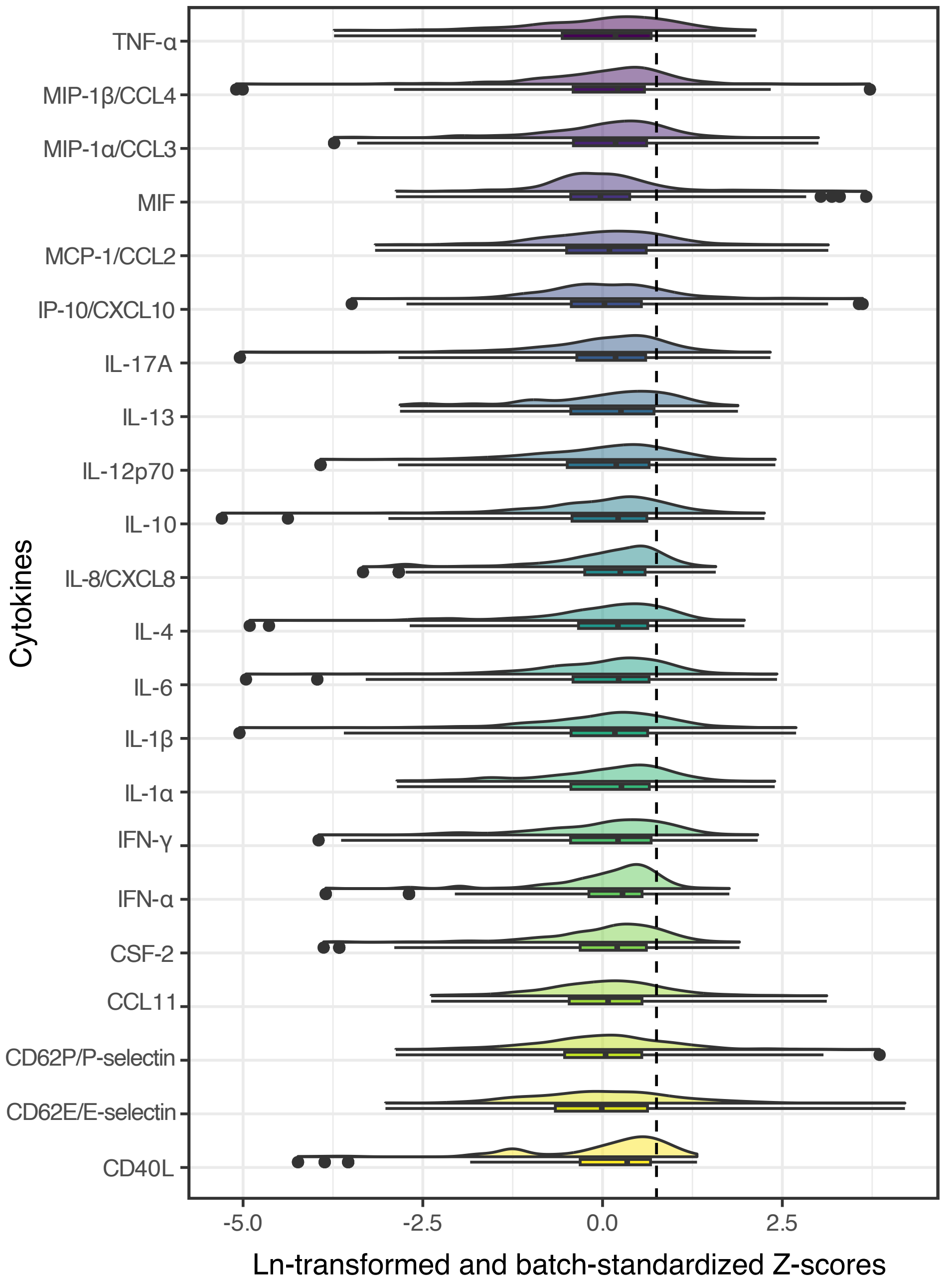


**Supplementary Figure S1. Boxplot and density of the baseline plasma levels of the 22 quantified cytokines.** The X-axis represents the Ln-transformed and batch-standardized cytokine levels (Z-scores), while the Y-axis displays the corresponding cytokines. The boxplots illustrate the interquartile range (IQR; 25th to 75th percentiles), with the line within each box indicating the median.

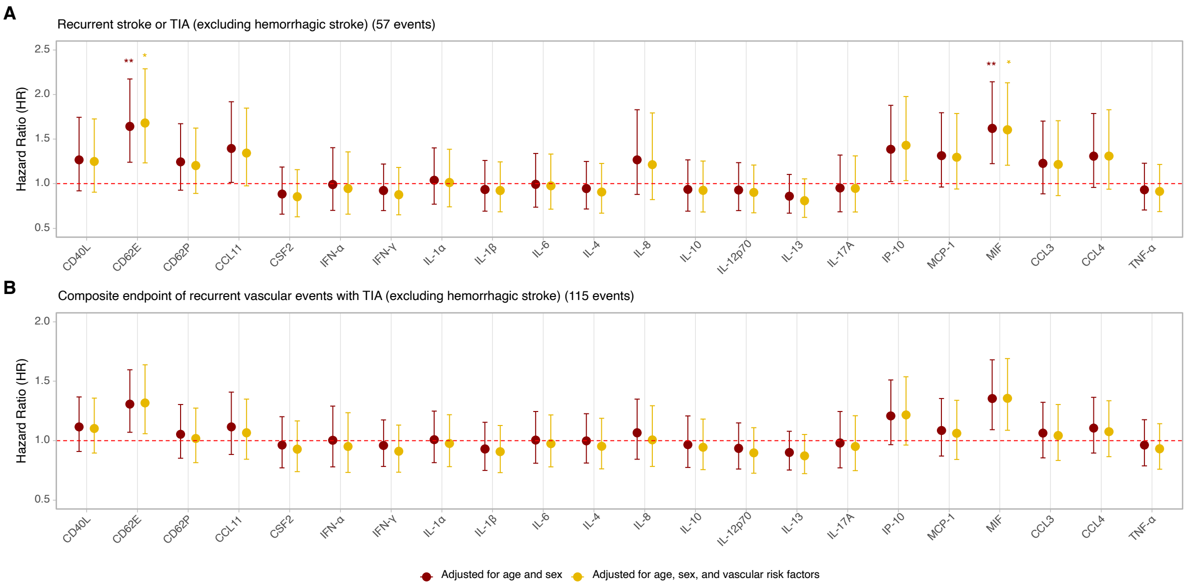


**Supplementary Figure S2. Associations between cytokine levels and recurrent stroke/TIA and recurrent vascular events (stroke, acute coronary syndrome, new-onset heart failure, or death) over the 5-year follow-up across different adjustment models in ischemic stroke patients**. Forest plots of hazard ratio (HR) for the associations of circulating baseline cytokine levels (per standard deviation increment in ln-transformed values) with **(A)** recurrent stroke/TIA (n=57) and **(B)** the composite endpoint of recurrent vascular events with TIA (n=115) over the 5-year follow-up period. The HR in the first model (red) are adjusted for age and sex, while those in the second model (yellow) are further adjusted for vascular risk factors, including hypertension, diabetes, current smoking, history of stroke, history of atrial fibrillation, stroke subtypes, anticoagulants, antihypertensive medications, antiplatelet agents, statins, and low-density lipoprotein levels. The vertical error lines indicate the 95% CI for each HR. Significance is indicated by *p-value < .05, **p-value < .01, representing FDR-corrected p-values for multiple comparisons. Abbreviations: TIA, transient ischemic attack; HR, hazard ratio; CI, confidence interval; FDR, false discovery rate.

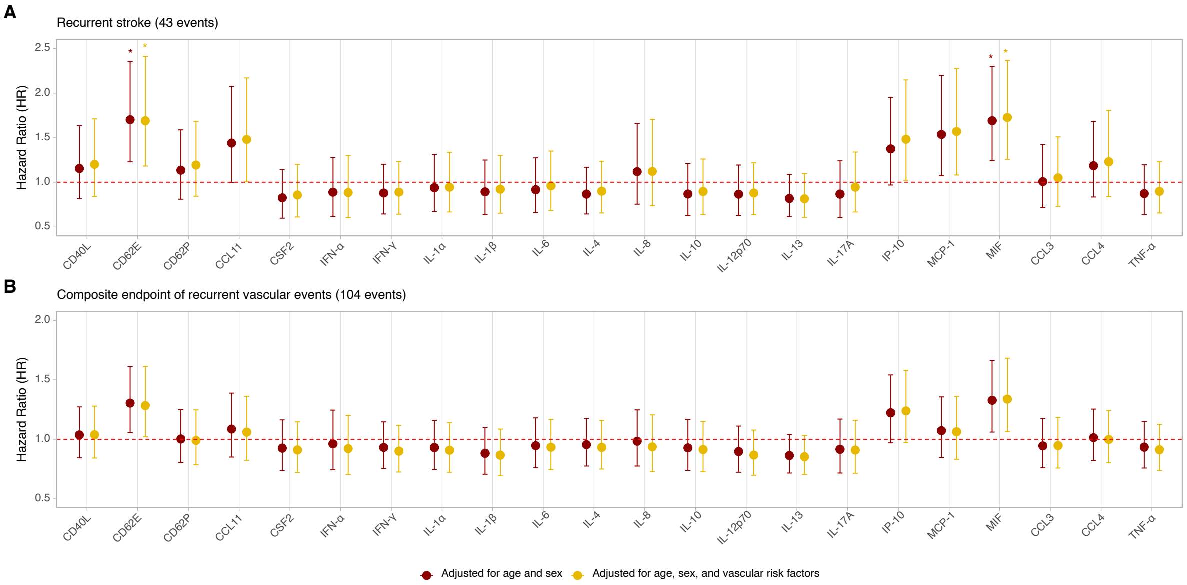


**Supplementary Figure S3. Associations of cytokine levels with recurrent stroke without TIA and recurrent vascular events without TIA (stroke, acute coronary syndrome, new onset of heart failure, or death) over the 5-year follow-up period**. Forest plots of hazard ratio (HR) for the associations of circulating baseline cytokine levels (per standard deviation increment in ln-transformed values) with **(A)** recurrent stroke (n=43) and **(B)** the composite endpoint of recurrent vascular events without TIA (n=104) over the 5-year follow-up period. The HR in the first model (red) are adjusted for age and sex, while those in the second model (yellow) are further adjusted for vascular risk factors, including hypertension, diabetes, current smoking, history of stroke, history of atrial fibrillation, stroke subtypes, anticoagulants, antihypertensive medications, antiplatelet agents, statins, and low-density lipoprotein levels. The vertical error lines indicate the 95% CI for each HR. Significance is indicated by *p-value < .05, representing FDR-corrected p-values for multiple comparisons. Abbreviations: TIA, transient ischemic attack; HR, hazard ratio; CI, confidence interval; FDR, false discovery rate.

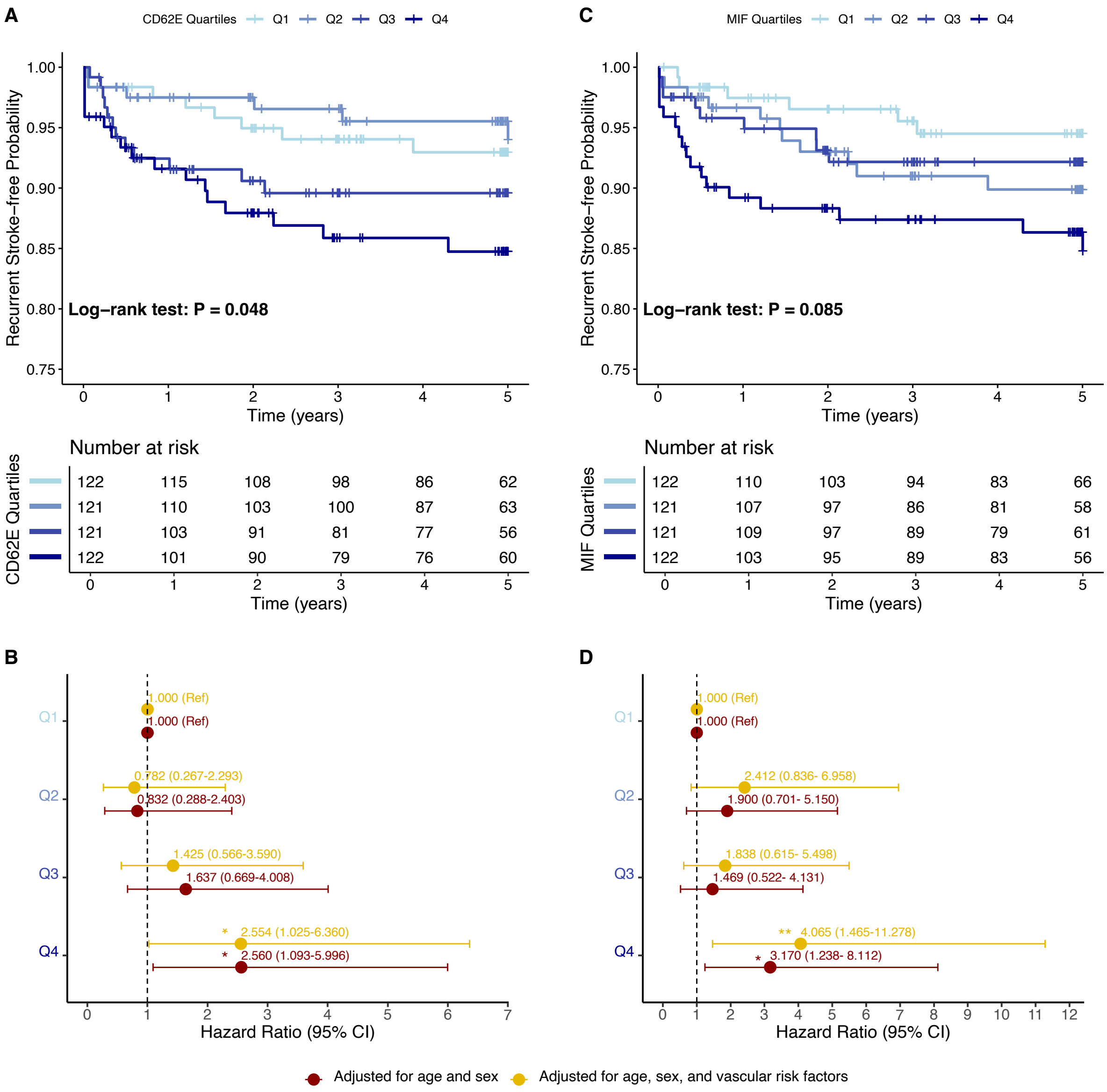


**Supplementary Figure S4. Dose-response associations of baseline CD62E and MIF levels with 5-year risk of recurrent stroke without TIA**. **(A)** Kaplan-Meier 5-year recurrent stroke-free survival curves for DEMDAS participants across quartiles (Q1 to Q4) of baseline circulating CD62E levels. **(B)** Forest plot of adjusted HRs for recurrent stroke across quartiles of baseline circulating CD62E levels. **(C)** Kaplan-Meier 5-year recurrent stroke-free survival curves for DEMDAS participants across quartiles (Q1 to Q4) of baseline circulating MIF levels. **(D)** Forest plot of adjusted HR for recurrent stroke across quartiles of baseline circulating MIF levels. In panels B and D, the HR in the first model (red) are adjusted for age and sex, while the HR in the second model (yellow) are further adjusted for vascular risk factors, including hypertension, diabetes, current smoking, history of stroke, history of atrial fibrillation, stroke subtypes, anticoagulants, antihypertensive medications, antiplatelet agents, statins, and low-density lipoprotein levels. Horizontal lines represent the 95% CI for each HR. Statistical significance is indicated by *p-value < .05, **p-value < .01. Abbreviations: TIA, transient ischemic attack; HR, hazard ratio; CI, confidence interval.

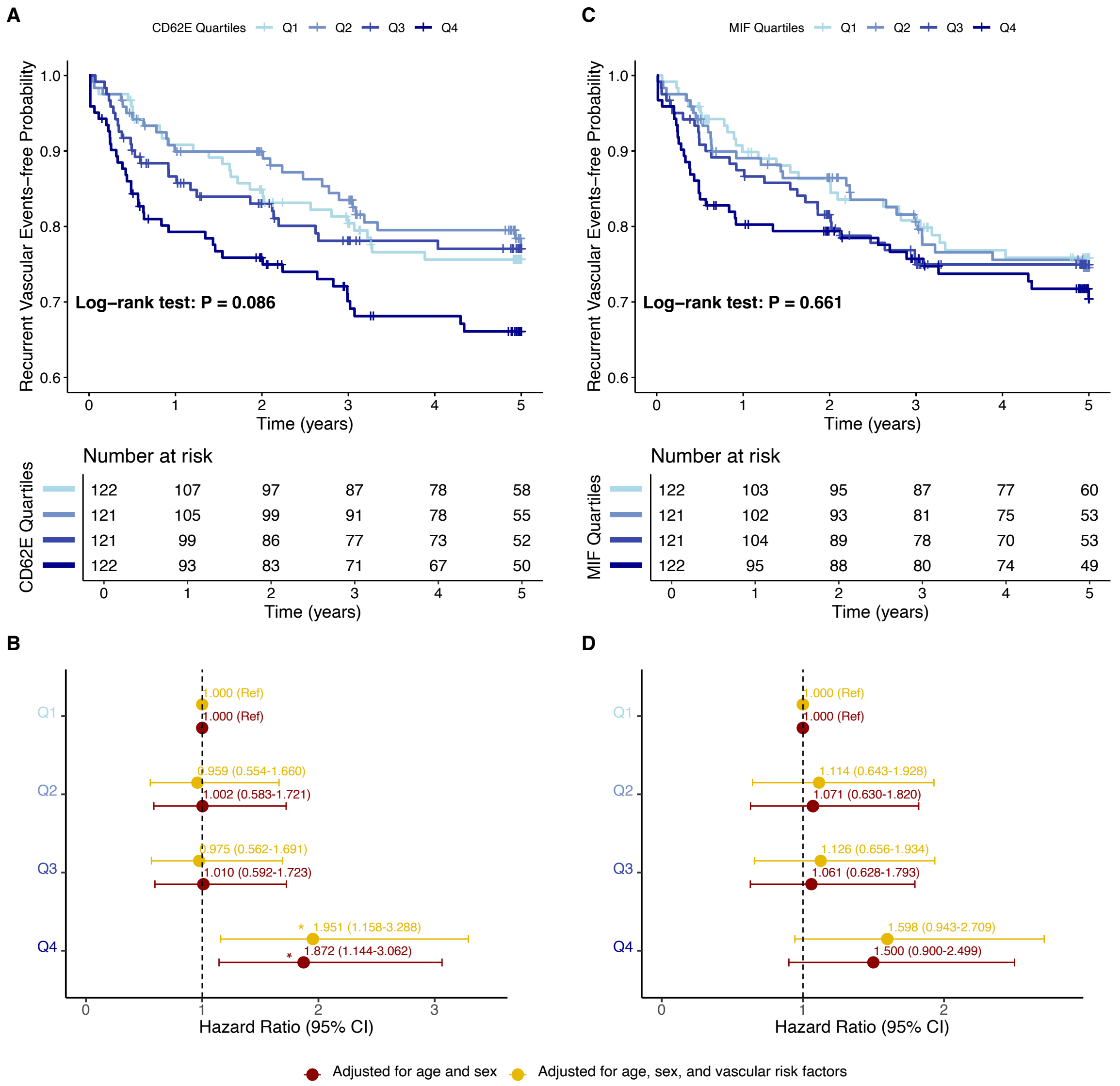


**Supplementary Figure S5. Dose-response associations of baseline CD62E and MIF levels with 5-year risk of recurrent vascular events (stroke, TIA, acute coronary syndrome, new onset of heart failure, or death). (A)** Kaplan-Meier 5-year recurrent vascular events-free survival curves for DEMDAS participants across quartiles (Q1 to Q4) of baseline circulating CD62E levels. **(B)** Forest plot of adjusted HR for recurrent vascular events across quartiles of baseline circulating CD62E levels. **(C)** Kaplan-Meier 5-year recurrent vascular events-free survival curves for DEMDAS participants across quartiles (Q1 to Q4) of baseline circulating MIF levels. **(D)** Forest plot of adjusted HR for recurrent vascular events across quartiles of baseline circulating MIF levels. In panels **(B)** and **(D)**, the HR in the first model (red) are adjusted for age and sex, while the HR in the second model (yellow) are further adjusted for vascular risk factors, including hypertension, diabetes, current smoking, history of stroke, history of atrial fibrillation, stroke subtypes, anticoagulants, antihypertensive medications, antiplatelet agents, statins, and low-density lipoprotein levels. Horizontal lines represent the 95% CI for each HR. Significance is indicated by *p-value< .05. Abbreviations: TIA, transient ischemic attack; HR, hazard ratio; CI, confidence interval.
